# Constructing the brief Diagnostic Criteria for Temporomandibular Disorders (bDC/TMD)

**DOI:** 10.1101/2023.08.29.23294531

**Authors:** Justin Durham, Richard Ohrbach, Lene Baad-Hansen, Stephen Davies, Antoon De Laat, Daniela Godoi Goncalves, Valeria V Gordan, Jean-Paul Goulet, Birgitta Häggman-Henrikson, Michael Horton, INfORM, Michail Koutris, Alan Law, Thomas List, Frank Lobbezoo, Ambra Michelotti, Donald R. Nixdorf, Juan Fernando Oyarzo, Chris Peck, Chris Penlington, Karen G. Raphael, Vivian Santiago, Sonia Sharma, Peter Svensson, Corine M. Visscher, Imamura Yoshiki, Per Alstergren

## Abstract

**Background:** Despite advances in Temporomandibular disorders’ (TMDs) diagnosis, the diagnostic process continues to be problematic in non-specialist settings.

**Objective:** To complete a Delphi process to shorten the Diagnostic Criteria for TMD (DC/TMD) to a brief DC/TMD (bDC/TMD) for the diagnoses with the most utility in general dentistry settings.

**Methods:** A international Delphi panel was created with 23 clinicians representing major specialities, general dentistry, and related fields. The process comprised a full day workshop, four virtual meetings, six rounds of electronic discussion, and finally an open consultation at a virtual international symposium.

**Results:** Within the physical axis (Axis 1) the self-report Symptom Questionnaire of the DC/TMD did not require shortening from 14 items for the bDC/TMD. The compulsory use of the TMD pain screener was removed reducing the total number of Axis 1 items by 18%. The DC/TMD Axis 1 10-section examination protocol (25 movements, up to 12 sets of bilateral palpations) was reduced to 4 sections in the bDC/TMD protocol involving 3 movements and 3 sets of palpations. Axis I then resulted in two groups of diagnoses: painful TMD (inclusive of secondary headache), and common joint-related TMD with functional implications. The Psychosocial Axis (Axis 2) was shortened to an ultra-brief 11 item assessment.

**Conclusion:** The bDC/TMD represents a substantially reduced and likely expedited method to establish (grouping) diagnoses in TMDs. This may provide greater utility for settings requiring less granular diagnoses for the implementation of initial treatment, for example non-specialist general dental practice.

## Introduction

The Diagnostic Criteria for Temporomandibular Disorders (DC/TMD), published in 2014, was designed to be used both in research as well as in general and specialist-level clinical settings ^1^. The DC/TMD, emerging from the Research Diagnostic Criteria for TMD (RDC/TMD) and based on data from the RDC/TMD Validation Project, represent the second generation of TMD diagnostic protocols ^2–4^. The dual axes approach in both protocols has been demonstrated to be both reliable and valid ^2–4^. Observation suggests that the revised and streamlined second axis for assessing the psychosocial domain has been particularly successful in achieving greater utilisation across specialist settings through its screening and comprehensive instrument selections. Overall, the DC/TMD is the currently recognized reference standard for comprehensive evaluation and classification of the most prevalent types of TMD ^5^.

The development of the DC/TMD was facilitated by the International Network for Orofacial Pain & Related Disorders Methodology (INfORM), a scientific group within the International Association for Dental Research (IADR). Prior to the publication of the DC/TMD, INfORM facilitated the implementation of field trials in both research and clinical settings. The Orofacial Pain: Prospective Evaluation and Risk Assessment (OPPERA) study, ^6^ comprised four study sites and took place over 12 years. OPPERA trained multiple examiners according to both pre-DC/TMD standards and published DC/TMD standards. Its annual examiner calibration and reliability assessment consistently yielded inter-examiner reliability 0.82 – 1.0 ^7^. The DC/TMD was also introduced immediately in another research setting, with annual examiner reliability assessments demonstrating its consistency in use ^8^. In clinical settings, multiple implementations of the DC/TMD were conducted (Buffalo, NY; Malmö, Sweden; Amsterdam, the Netherlands; and other settings), demonstrating the applicability of training across the profession ^9, 10^.

INfORM appoints DC/TMD Training and Reliability Assessment Centres (Malmo, Sweden; Umeå, Sweden; Aarhus, Denmark; Oulu, Finland; Leipzig, Germany; and Tokyo, Japan) to disseminate DC/TMD to clinicians and researchers by providing structured courses in the DC/TMD and a structured reliability assessment procedure. These centres have provided numerous lectures and courses for non-specialist dentists. Collectively, these efforts have demonstrated that the painful TMDs phenotype can be identified in differing settings with very high reliability and validity, equal to that of any type of medical disorder. Whilst pain is inherently a subjective experience, the clinical correlates of having pain can be evaluated with sufficient rigor. A notable consequence is that a more accurate and reliable diagnosis is likely to minimize inappropriate and unneeded therapies and (consequently) lead to better prognosis and treatment.

Despite numerous international efforts to implement the DC/TMD since its publication, the use of the DC/TMD in non-specialist settings in dentistry has remained disappointingly low^11, 12^. This low implementation seems to have been due to three major obstacles: i) the amount of training necessary to adequately use DC/TMD; ii) the belief that clinical implementation of the physical examination is excessively time-consuming; and iii) the complexity of implementing the psychosocial axis (Axis 2) ^13^. A simplified and brief version of the DC/TMD can address, at least in part, these three obstacles.

Supportive evidence for the viability of a simplified brief version comes from two evaluations of the DC/TMD. First, Swedish clinicians who self-taught the DC/TMD from video training materials achieved comparable diagnoses to those made by clinicians who were formally taught the DC/TMD through a two-day theoretical and practical DC/TMD course, when compared to diagnoses from the reference standard examiner ^10^. Second, the use of mandatory commands, in comparison to simpler non-standardised language targeting the operational goal of each procedure, did not improve sensitivity and specificity for the DC/TMD diagnoses myalgia, arthralgia, headache attributed to TMD, and degenerative joint disease, which are the important diagnoses in non-specialist settings ^9^. While the latter was specific to Swedish language, it may have implications elsewhere. Agreement was excellent between DC/TMD examination conducted with the aid of video communication technology and a reference standard DC/TMD in-person examination for the diagnoses of myalgia and arthralgia but not for the diagnosis of disk displacement with reduction ^14^. Remote assessment of patients with pain-related TMDs using the DC/TMD is, therefore also feasible and presents a high degree of accuracy.

The use of a simplified and brief DC/TMD in primary settings would still entail use of the dual axis approach within the DC/TMD: Axis 1, which leads to physical diagnosis; and Axis 2, which provides psychosocial assessment of the person. Within Axis 1 there is currently: a self-report screener for painful TMDs ^15^ consisting of three items; a self-report symptom questionnaire consisting of 14 items that is used as part of the diagnostic algorithms; and a physical examination requiring a 10-section protocol to be performed and recorded.

Altogether the complete DC/TMD physical examination 10-section protocol requires 25 movements and up to 12 sets of palpations to be performed on each side separately of the muscles and joint (‘bilateral palpations’). An experienced clinician can conduct this examination on most individuals with pain complaints in about 5 minutes by tailoring some of the more time-consuming procedures to the chief complaint and using active decision-making during the examination procedure itself ^16^. This approach to using the DC/TMD framework, however, is not typically possible in most primary care settings where clinicians may be less experienced.

Within Axis 2, both brief screening and comprehensive screening approaches for patient assessment via the self-report instruments are described in the DC/TMD ^1^. For each of the instruments used within these approaches, specific purposes relevant to pain conditions and management have also been described ^17^. An ultra-brief screening approach to Axis 2 has also been identified, which captures substantial relevant information coupled with reduced patient and clinician burden ^18^. The elements of Axis 2, as well as the three approaches to its use, are demonstrated in Table 1.

**Table 1.**
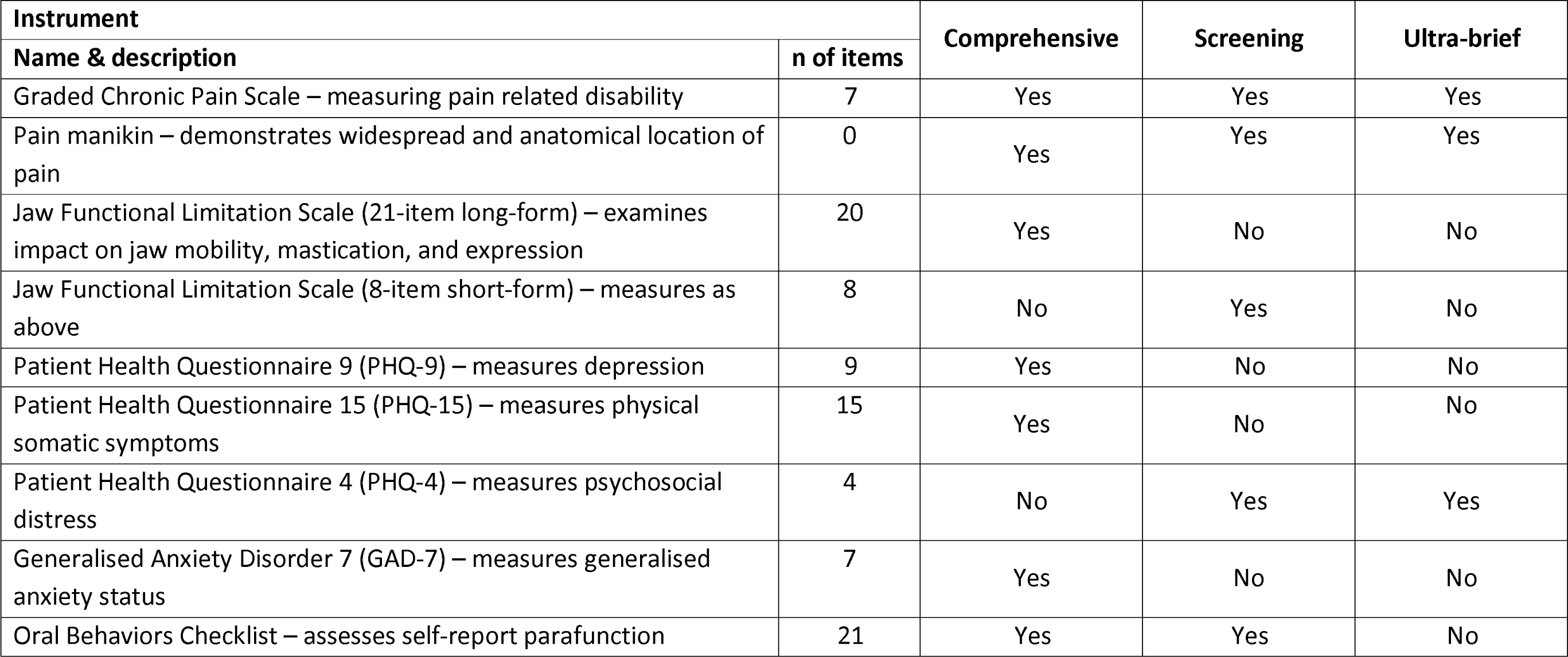
Axis 2 comprehensive, screening and ultra-brief axis contents.

The DC/TMD has received a large amount of attention worldwide, resulting in translations into more than 20 languages across as many countries. The 2014 publication is also one of the top 100 highly cited papers in dentistry ^19^. Despite this, the implementation of the DC/TMD in settings outside of orofacial pain (specialist) clinics is low even though it was designed for applicability to *both* research and routine clinical practice. This is particularly true within time-pressurised healthcare systems and non-specialist, community-based general dental practitioner settings. Recognizing this limitation, INfORM elected to initiate a Delphi process to refine the DC/TMD to the briefest criteria with clinical utility and create the brief DC/TMD, or ‘bDC/TMD’.

The aim of this study was therefore to complete a Delphi process to shorten the DC/TMD to a bDC/TMD for the diagnoses with greatest utility in general dentistry settings.

## Methods

A full day Delphi workshop (7 hours excluding breaks) was hosted and facilitated by INfORM at the International Association of Dental Research’s (IADR) Annual Scientific Meeting in London, UK in 2018.

Invitations were sent in advance of the workshop to colleagues practising in community-based primary general dental care and in specialist settings from around the world. Twenty-three individuals were invited (Supplementary information table 1) of which 21 were able to attend in person and the remaining 2 individuals contributed to subsequent discussions and drafting processes.

The workshop began with a structured introduction and discussion of the draft guiding principles for the reduction of the DC/TMD to a bDC/TMD. The draft version of these principles was then revised and endorsed by all workshop participants. The workshop then divided into two time-limited sessions: the morning session to consider Axis 1 of the DC/TMD (physical diagnoses); and the afternoon session to consider Axis 2 (psychosocial assessment). Participants were allocated to one of four groups for the duration of the workshop. Each of these groups contained 5-6 individuals and was led by an experienced facilitator (CV, FL, DN, CP) who helped ensure all voices/opinions were heard, considered, and discussed. Facilitators also kept notes from the discussions to which they and the principal investigators could refer. The principal investigators (JD, PA, RO) took field notes and circulated among groups to help ensure consistency.

An identical format was used for the morning and afternoon sessions:

- One of the principal investigators provided a brief structured introduction on the relevant material for that session, and accompanying DC/TMD documentation was available to all participants for reference throughout;
- Proposed changes from DC/TMD to bDC/TMD were discussed intra-group and moderated by the group facilitator, ensuring that any proposed revisions/reductions were made with reference to both the guiding principles for the new instrument and the current DC/TMD diagnostic decision trees and diagnostic criteria table (https://ubwp.buffalo.edu/rdc-tmdinternational/tmd-assessmentdiagnosis/dc-tmd/). In these discussions, Axis 1 construction was examined to reduce the items within Axis 1 to the smallest sub-set that would provide acceptable (grouping) diagnoses for: a) Painful TMDs: myalgia, headache attributed to TMD, arthralgia; and b) Common joint-related TMDs with implications for function: degenerative joint disease, disc displacement disorders, and subluxation. This was performed by each of the four groups independently against the full DC/TMD criteria;
- Each of the four groups provided summary presentations to the whole panel regarding proposed changes and their rationale.

Following the four group presentations, a plenary session with all participants, facilitated by the three principal investigators, allowed for full discussion of all proposals.

Following the completion of the workshop, the three principal investigators compiled and organized notes with regard to recurring themes of discussion and consensus achieved at that point. They used these data to construct and present the initial draft instrument (bDC/TMD) and the conclusions drawn from the workshop to the workshop participants in order to triangulate and validate the instrument’s content and the interim conclusions. This process involved a total of 6 rounds of email discussion and 4 virtual meetings. There was then a wider consultation process using a virtual open-invitation symposium held at IADR’s annual scientific meeting in 2022 to invite feedback from any individual interested in commenting on the draft instrument.

## Results

For the purposes of clarity, the results are presented in three sections followed by a summary of the final content of the bDC/TMD protocol.

### Guiding principles for the bDC/TMD protocol

The draft principles for the protocol presented to the Delphi panel were:

1. applicability to non-specialist care settings must maintain both axes but especially in relation to Axis 2 ^12^;
2. for Axis I, the DC/TMD sensitivity and specificity should be retained as far as possible for the painful TMDs diagnoses (myalgia, arthralgia, headache attributed to TMD) and the other common joint-related TMDs with a focus on functional implications (degenerative joint disease, subluxation, acute closed lock and a collapsed category of other disc based TMDs);
3. the possibility to learn, at distance without the need to attend for face-to-face training, the processes involved in using the bDC/TMD.

The discussion by all participants around the first principle of applicability to non-specialist care related to two parameters of the final protocol. The first parameter was the time required to complete the physical diagnosis assessment in clinic, and the second was the level of diagnostic information and sub-typing provided by completion of both axes of the protocol for the non-specialist.

The Delphi panel discussed the time available in a non-specialist clinic to complete the documentation associated with the protocol and the physical examination. The panel agreed unanimously that utilisation of self-report must be optimised and preferably maximised to save the clinician time in clinic. Correspondingly, the group felt that the time required in the clinic to complete the full bDC/TMD, that is the physical examination and processing any self-report outcomes, should be no more than 10 minutes. The group suggested that optimisation of self-report could be achieved through two main methods dependent on the infrastructure and expertise available to the clinician: dental assistant/nurse generating outcomes from self-report instrumentation ahead of clinic or during the history taking; and online administration of the self-report instrumentation ahead of clinic through a secure web-portal.

The panel explored and discussed the level of diagnostic information that could be potentially achieved from the 12 most frequently presenting sub-types of TMDs as implemented in the DC/TMD, versus what could be potentially achieved by using simple overarching grouping diagnoses of painful TMDs and of common joint-related TMDs with implications for function. Despite appreciating the benefits that a more specific sub-type diagnosis could bring for research and specialist care purposes by use of the DC/TMD, the Delphi participants felt that the state of practice in non-specialist care meant there was little to be gained from the bDC/TMD leading to all 12 sub-type diagnoses, especially as evidence-based initial management would not differ ^20, 21^. Moreover, the intent of the bDC/TMD is to yield a clinically expedient initial grouping diagnosis, not diagnostic information with the necessary detail required for research or specialist settings. A related recommendation was that if the bDC/TMD was successfully incorporated into routine non-specialist care that there should be a further review and revision of this position in future years – which was arbitrarily set at 10 years after publication of this paper.

The panel therefore elected to set the outputs from both axes of the bDC/TMD at the simplest possible level with overarching (grouping) diagnoses of painful TMDs (myalgia, arthralgia, headache attributed to TMD) or common joint-related TMDs with implications for function (degenerative joint disease, subluxation, acute closed lock, and a collapsed category of other disc based TMDs). This was based first on the fact that, firstly, this is a step forward from the current position of general clinical practice tending towards the use of “TMD” as a singular “catch-all” term. Secondly, in non-specialist practice there would be very little difference between the management of the 12 most frequently presenting sub-types of TMDs, but explanations of the sources of pain or limitation would differ, and education of patients and clinicians may improve as a result. The same was felt to be true of outputs from Axis 2 where an ultra-brief screening was sought to identify any comorbidities of importance that might require further exploration or care from more specialised services.

The group examined the recent data available from the literature on the use of self-education via video training for the current DC/TMD. They accepted the trade-off between the potential limitations in skill acquisition and skill retention with self-education versus its benefits of ease and accessibility over formally delivered training at nominated centres. As a result, they came to a consensus that video instructions would suffice for bDC/TMD training especially given that the intention was to produce a grouping diagnosis rather than specific sub-typing of TMDs into the 12 common types.

In summary, therefore, the draft guiding principles were revised and agreed upon within the workshop as:

1. The bDC/TMD axis 1 and axis 2 should take no more than 10 minutes of provider time to complete in its entirety in clinic, and the structure of the bDC/TMD should optimise and maximise the patient’s use of self-report instrumentation;
2. The bDC/TMD should produce Axis 1 grouping diagnoses or combinations thereof with clinical utility rather than down to the level of all 12 of the most frequently presenting sub-types of TMDs. These groupings are painful TMDs (inclusive of myalgia, arthralgia, and headache attributed to TMD) or other common joint-related TMDs with implications for function (inclusive of degenerative joint disease, subluxation, acute closed lock, and a collapsed category of other disc based TMDs). Axis 2 should focus on an ultra-brief screen to demonstrate yellow or red flags in psychosocial status ^22^;
3. Training for clinicians in the use of the bDC/TMD will be provided via distance learning such as through video training material. The training component will require additional resources to effectively implement across languages and settings, but it is deemed an immediately achievable goal that can be empirically tested later.

### Axis 1

The guiding principles informed the assessment and reduction of the Axis 1 content of the DC/TMD: the TMDs pain screener, the patient Symptom Questionnaire and examination sections. The reduction was discussed in groups and the whole panel then agreed on the reductions.

Table 2 demonstrates both the Symptom Questionnaire items and the panel’s decision and rationale on their retention or removal in the bDC/TMD. The panel elected to remove the TMDs Pain Screener as compulsory from the bDC/TMD protocol in order to allow clinicians and healthcare systems around the world to implement their preferred screening tool(s) for TMDs, given that screening is often context and setting dependent. This resulted in a three item reduction for the total number of items within the self-report instruments for Axis 1. The panel identified 3 further items (Table 2) to remove from the Symptom Questionnaire that, on initial inspection, were not required within the envisaged revised diagnostic decision-tree for the new bDC/TMD that would utilize group-level diagnoses. However, ensuring that the prevalent patient complaints were adequately captured by the SQ as well as initial testing of a bDC/TMD diagnostic algorithm resulted in retaining the full DC/TMD Symptom Questionnaire. Consequently, the self-administered Axis 1 portion of the bDC/TMD consists of the existing Symptom Questionnaire (containing 14 items) yet representing a 18% reduction in self-administered items in Axis 1 compared to the original parent instruments in the DC/TMD (total 17 items across screener and symptom questionnaire – supplementary information “bDC/TMD self-report SQ and Axis 2”).

**Table 2.**
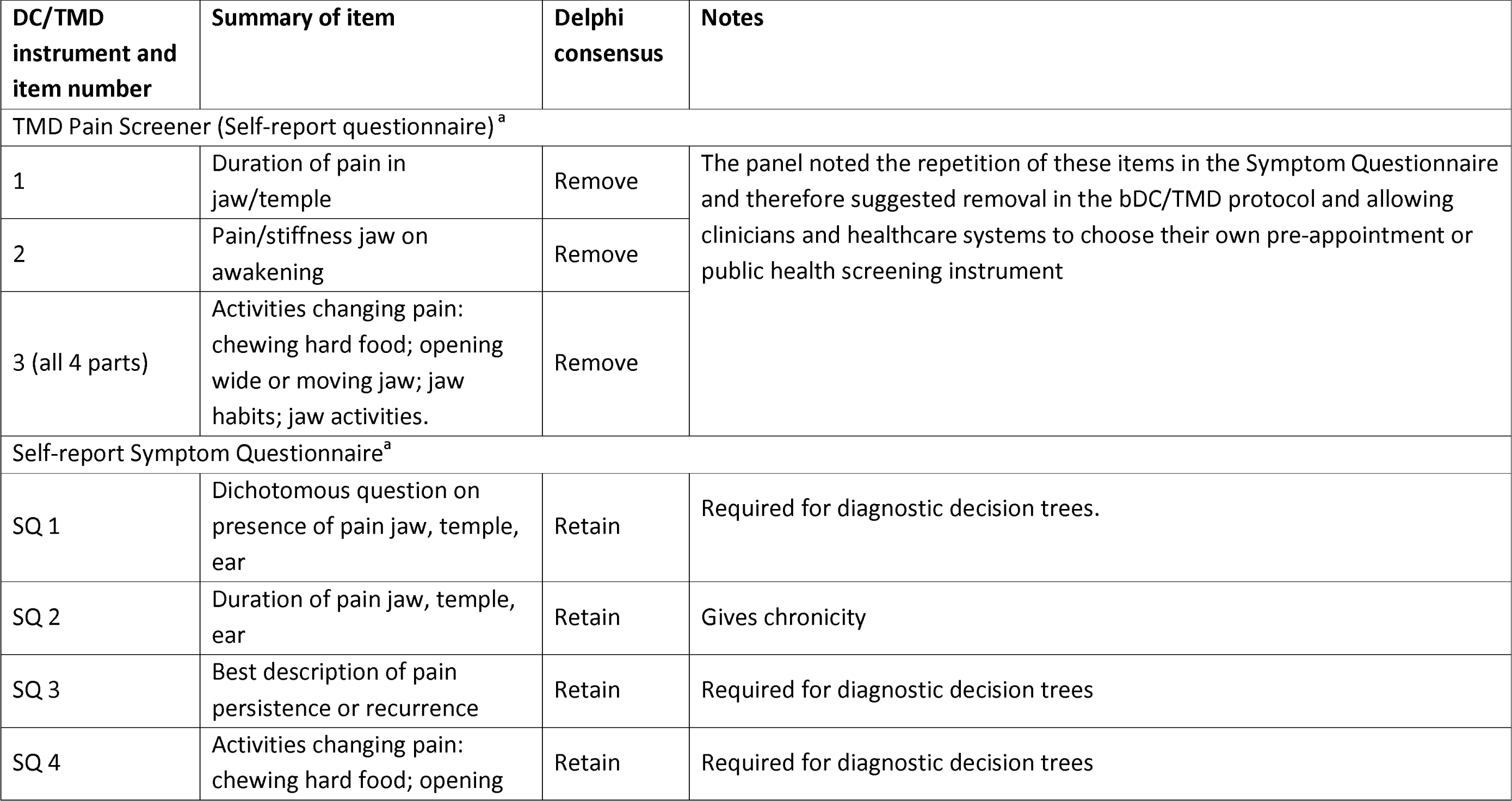
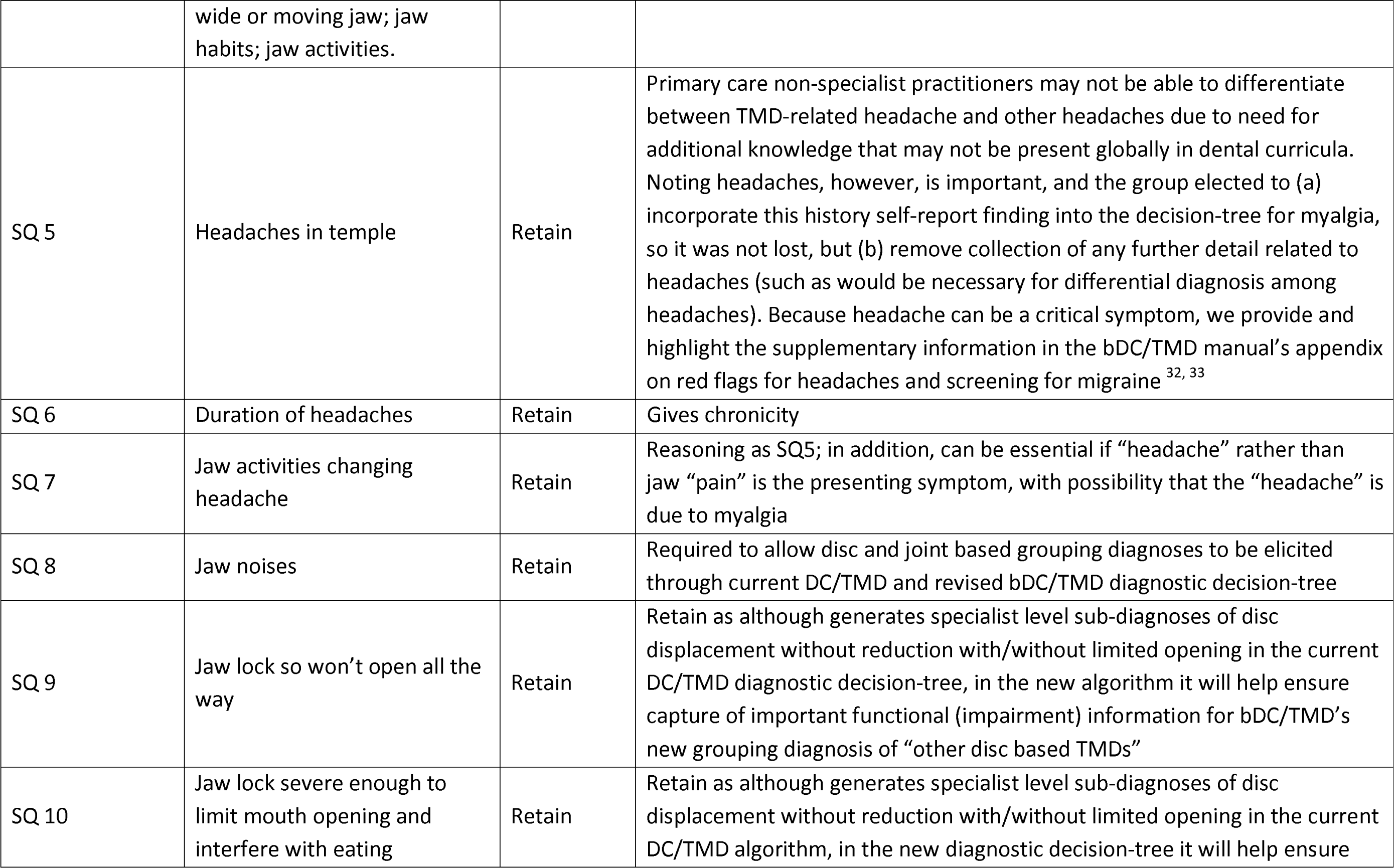
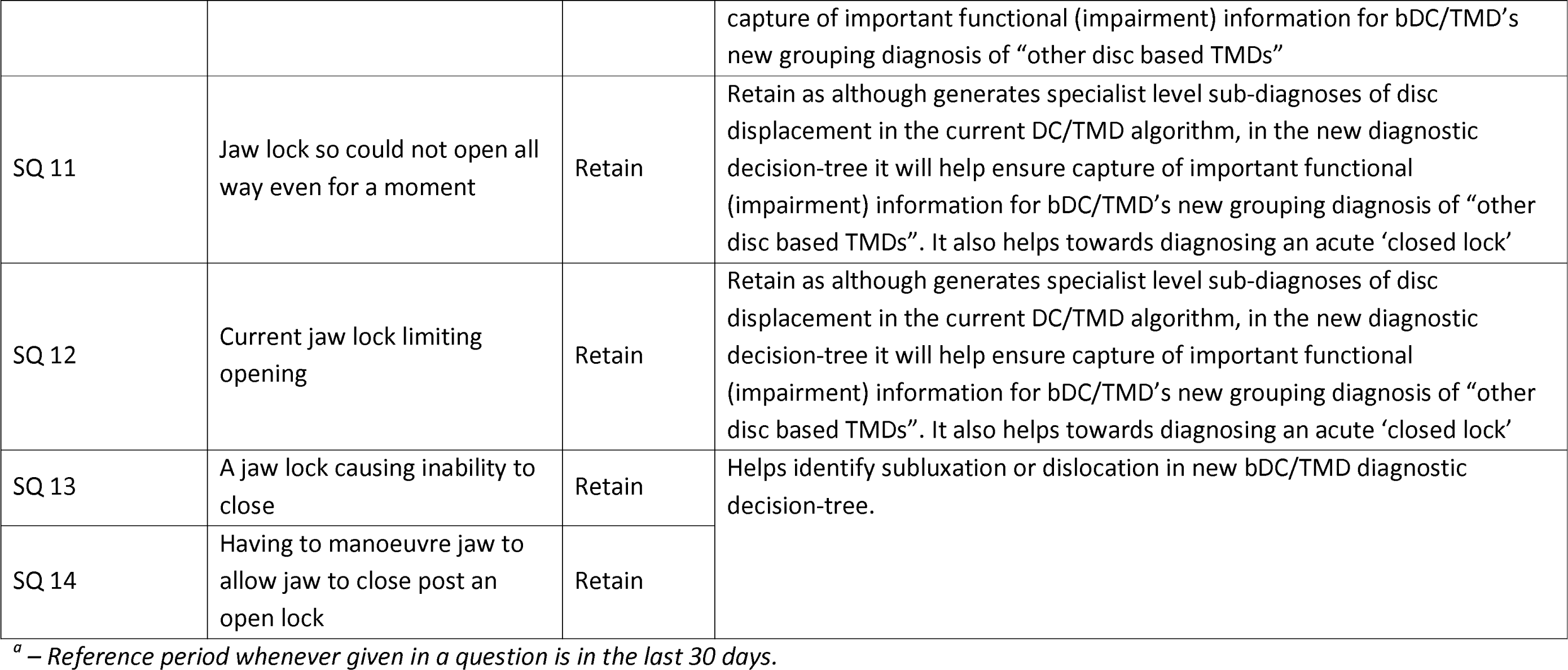
Original DC/TMD Axis 1 TMD Pain Screener and Symptom Questionnaire items and reduction of these items by Delphi panel.

| DC/TMD instrument and item number | Summary of item | Delphi consensus | Notes |
| --- | --- | --- | --- |
| TMD Pain Screener (Self-report questionnaire) <sup>a</sup> |  |  |  |
| 1 | Duration of pain in jaw/temple | Remove | The panel noted the repetition of these items in the Symptom Questionnaire and therefore suggested removal in the bDC/TMD protocol and allowing clinicians and healthcare systems to choose their own pre-appointment or public health screening instrument |
| 2 | Pain/stiffness jaw on awakening | Remove |  |
| 3 (all 4 parts) | Activities changing pain: chewing hard food; opening wide or moving jaw; jaw habits; jaw activities. | Remove |  |
| Self-report Symptom Questionnaire <sup>a</sup> |  |  |  |
| SQ 1 | Dichotomous question on presence of pain jaw, temple, ear | Retain | Required for diagnostic decision trees. |
| SQ 2 | Duration of pain jaw, temple, ear | Retain | Gives chronicity |
| SQ 3 | Best description of pain persistence or recurrence | Retain | Required for diagnostic decision trees |
| SQ 4 | Activities changing pain: chewing hard food; opening wide or moving jaw; jaw habits; jaw activities. | Retain | Required for diagnostic decision trees |
| SQ 5 | Headaches in temple | Retain | Primary care non-specialist practitioners may not be able to differentiate between TMD-related headache and other headaches due to need for additional knowledge that may not be present globally in dental curricula. Noting headaches, however, is important, and the group elected to (a) incorporate this history self-report finding into the decision-tree for myalgia, so it was not lost, but (b) remove collection of any further detail related to headaches (such as would be necessary for differential diagnosis among headaches). Because headache can be a critical symptom, we provide and highlight the supplementary information in the bDC/TMD manual's appendix on red flags for headaches and screening for migraine<sup>32, 33</sup> |
| SQ 6 | Duration of headaches | Retain | Gives chronicity |
| SQ 7 | Jaw activities changing headache | Retain | Reasoning as SQ5; in addition, can be essential if "headache" rather than jaw "pain" is the presenting symptom, with possibility that the "headache" is due to myalgia |
| SQ 8 | Jaw noises | Retain | Required to allow disc and joint based grouping diagnoses to be elicited through current DC/TMD and revised bDC/TMD diagnostic decision-tree |
| SQ 9 | Jaw lock so won't open all the way | Retain | Retain as although generates specialist level sub-diagnoses of disc displacement without reduction with/without limited opening in the current DC/TMD diagnostic decision-tree, in the new algorithm it will help ensure capture of important functional (impairment) information for bDC/TMD's new grouping diagnosis of "other disc based TMDs" |
| SQ 10 | Jaw lock severe enough to limit mouth opening and interfere with eating | Retain | Retain as although generates specialist level sub-diagnoses of disc displacement without reduction with/without limited opening in the current DC/TMD algorithm, in the new diagnostic decision-tree it will help ensure capture of important functional (impairment) information for bDC/TMD's new grouping diagnosis of "other disc based TMDs" |
| SQ 11 | Jaw lock so could not open all way even for a moment | Retain | Retain as although generates specialist level sub-diagnoses of disc displacement in the current DC/TMD algorithm, in the new diagnostic decision-tree it will help ensure capture of important functional (impairment) information for bDC/TMD's new grouping diagnosis of "other disc based TMDs". It also helps towards diagnosing an acute 'closed lock' |
| SQ 12 | Current jaw lock limiting opening | Retain | Retain as although generates specialist level sub-diagnoses of disc displacement in the current DC/TMD algorithm, in the new diagnostic decision-tree it will help ensure capture of important functional (impairment) information for bDC/TMD's new grouping diagnosis of "other disc based TMDs". It also helps towards diagnosing an acute 'closed lock' |
| SQ 13 | A jaw lock causing inability to close | Retain | Helps identify subluxation or dislocation in new bDC/TMD diagnostic decision-tree. |
| SQ 14 | Having to manoeuvre jaw to allow jaw to close post an open lock | Retain |  |

<sup>a</sup> – Reference period whenever given in a question is in the last 30 days.

The standardised DC/TMD examination sections and procedures, as reflected in the examination form (available at https://ubwp.buffalo.edu/rdc-tmdinternational/tmd-assessmentdiagnosis/dc-tmd/ and summarised in Table 3), were evaluated, and the panel identified examination procedures to remove or revise as described in Table 3. These revisions resulted in the bDC/TMD consisting of 4 examination sections which involve 3 movements and 3 sets of palpations to perform and record. In total, this represents a 79% reduction in performing/recording in the bDC/TMD compared to the parent instrument (DC/TMD: 10 sections involving 25 movements and 12 sets of palpations). They also resulted in a simplification of the decision trees for diagnosis (Supplementary information – “Decision trees for bDC/TMD”).

**Table 3.** Original DC/TMD Axis 1 examination items and agreed reduction of these items by Delphi panel.

| DC/TMD examination item number<sup>a</sup> | Summary of examination item | Delphi consensus | Notes |
| --- | --- | --- | --- |
| E1a | Patient's identification of areas of pain. Areas of interest are demonstrated by light touch by examiner as: Temporalis, Masseter, TMJ, other masticatory muscles, non-masticatory structures | Retain and revise | Remove other masticatory muscles and non-masticatory structures as not incorporated within new bDC/TMD diagnostic decision-tree producing broad grouping diagnoses |
| E1b | Anatomical area affected by headache | Remove | Not required for new bDC/TMD diagnostic decision tree |
| E2 | Incisal relationship | Remove | Remove as additional detail unrequired for broad grouping diagnoses in new bDC/TMD diagnostic decision-tree |
| E3 | Opening pattern | Remove | Already considered 'supplemental' in DC/TMD largely due to poor reliability of this assessment; not used by its diagnostic decision-tree |
| E4 | Opening movements | Retain and revise | Simplify to only pain-free and maximum unassisted opening given (a) this differentiates between painful and non-painful TMDs and (b) helps with defining acute closed lock (disc displacement without reduction with limited opening). Reduce to 3 examination steps for a combined examination of E4, E6, E8 with 1 out of 3 movements being a positive finding with no further movement required once positive identified |
| E5 | Lateral movements | Remove | Have low reliability, contribute little to diagnosis, and difficult for patients to reproduce. |
| E6 | Joint noises on opening and closing | Retain and revise | Adjusted and shortened examination steps, as stated under E4 |
| E7 | Joint noises on lateral and protrusive movements | Remove | Low yield of positive results from these movements. |
| E8 | Joint locking | Retain and revise | Adjusted and shortened examination steps, as stated under E4 |
| E9 | Muscle and TMJ pain with palpation | Retain and revise | Remove reporting of palpation results from specified anatomical sub-sections of muscles (e.g., origin, body, insertion of masseter) as unnecessary level of detail for bDC/TMD diagnostic decision tree. The protocol will specify to focus examination on anterior temporalis and body of masseter as initial sites to examine with palpation, and to extend beyond those areas as needed, but form only requires findings at the muscle level.<br>Remove supplemental muscles from examination as non-contributory to diagnostic decision tree currently in DC/TMD and therefore unnecessary for bDC/TMD |

<sup>a</sup> – *Reference period whenever given in a question is in the last 30 days.* It is however important to note that whilst the 30-day period is useful and appropriate for most patients, sometimes a complaint may require that the reference frame be modified.

### Axis 2

The screening and comprehensive versions of the DC/TMD Axis 2 were reviewed by each group, and discussions on the content of the bDC/TMD’s Axis 2 were informed by both the guiding principles for the new instrument and the utility of the presently available instruments.

The panel then had substantial discussion about the simplest and most interpretable way of understanding the level of complexity of the patient presenting to the non-specialist practitioner. They were unanimous in their opinion regarding the clinical utility of the body pain manikin which was to be included in the bDC/TMD Axis 2. However, challenges with the further assessment of the patient’s psychosocial status were identified. These challenges included the complexity of calculation of the Graded Chronic Pain Scale (GCPS) and the fact that whilst the Patient Health Questionnaire-4 (PHQ-4) is ubiquitous in some healthcare systems it is not in others.

The complexity of the GCPS scoring was remediated by the agreement to change to the newer GCPS with 30-day reference period which has simpler scoring ^23^. The simpler scoring is based on only six items: three for pain intensity, and three for pain-related interference in functioning. Items dropped from the DC/TMD GCPS-30 days instrument are number of pain days in the prior six months and number of disability days due to facial pain in prior 30 days. The panel were unanimous in their appreciation of the utility of GCPS scores in helping assess pain intensity, pain-related disability and give an indication of complexity and prognosis. It was therefore suggested to form part of the bDC/TMD Axis 2 on the proviso the version with simpler scoring was used, which omits the scoring of disability days and grade of chronic pain which requires far more complex scoring ^23^.

The discussion about whether to include PHQ-4 as the standard assessment instrument for distress was related to two factors: it is not used by every healthcare system globally and there were at that time no data pertaining to its prognostic capabilities for orofacial pain. The final decision was to include PHQ-4 as a measure in the bDC/TMD for health care systems that use it or can use it, as it gives a good overview of the patient’s level of psychosocial distress and data were published on its prognostic capabilities over the time period of the Delphi process ^24^. For other settings it may be appropriate to substitute the PHQ-4 with a different distress screener.

The other elements of the screening and comprehensive versions of the Axis 2 instrument set were reviewed for whether they added to non-specialist patient care. These instruments included the Oral Behaviors Checklist, PHQ-9, PHQ-15, Jaw Functional Limitation Scale, and GAD-7. The information obtained from these instruments was regarded to add little to the initial management of an individual with a TMDs at the level of the non-specialist and were therefore omitted from the bDC/TMD’s proposed Axis 2.

The new proposed Axis 2 for bDC/TMD consists of 11 items (Supplementary information “bDC/TMD self-report SQ and Axis 2”) as compared to the original DC/TMD standard screening (40 items) and comprehensive (79 items) versions of Axis 2. All three instruments that provide the 11 items have been formally translated, to date, into 21 languages as part of the DC/TMD translation efforts. The 11 items independently selected by the Delphi panel for an ultra-brief screener for non-specialist care match those selected elsewhere for use with orthodontic patients (e.g., Ohrbach and Michelotti ^18^), attesting to the sensible value of these 11 items in terms of information gained vs patient burden.

### Summary of results and final instrument

In summary, the final agreed reduction of DC/TMD to bDC/TMD resulted in the instrument in the supplementary materials (Supplementary information “bDC/TMD self-report SQ and Axis 2”) which consists of:

– Axis 1

`o` Self-report symptom questionnaire (14 items)
`o` Examination form (total 4 individual examination sections)
– Axis 2

`o` Pain manikin (1 item)
`o` GCPS (up to 8 items but modified, for scoring purposes, to 6 with publication of Sharma et al ^23^)
`o` PHQ4 (4 items)

## Discussion

This study provides a first version of a bDC/TMD for use in non-specialist settings. It describes the multidisciplinary, international process for reducing the DC/TMD to a bDC/TMD. The bDC/TMD can now be disseminated to non-specialist settings for use. At the same time, it is important to gather data from studies or field tests of bDC/TMD to improve this version and identify potential gaps in its usefulness. For example, the revised graded chronic pain scale (GCPS-R, ^25^) might be examined against the current 1-month GCPS. It is also important to note that the bDC/TMD aims to characterise patients presenting with symptoms similar to TMDs or presumed TMDs due to having a positive result on a TMDs screening instrument such as the TMD Pain Screener ^15^ or 3Q/TMD ^26–28^ prior to consultation with the dental professional. It does not seek to rule out other diagnoses and neither does its parent instrument, the DC/TMD. There will however be information in the appendices to the manual for bDC/TMD on the expanded DC /TMD taxonomy ^29^, the international classification for orofacial pain ^30^, red flag symptoms for headache and for trismus ^31, 32^, and a validated migraine screening instrument ^33^ to help practitioners if they want to explore differential diagnoses.

The bDC/TMD represents a substantial reduction and simplification of examination items and decision trees (Supplementary information – “Decision trees for bDC/TMD”) required to assess in comparison to DC/TMD in addition to the removal of a 3-item compulsory TMD pain screener. Taken together with studies demonstrating that the DC/TMD mandatory commands and two-day training/calibration course may not be critical for non-specialist settings, the bDC/TMD should be implementable in general dental settings. It is likely that the diagnostic procedures for identifying painful TMDs and common joint-related TMDs with functional implications will have sufficient sensitivity and specificity compared to the respective reference standard diagnoses derived from the DC/TMD. Such optimism derives from careful consideration of how to best operationalize the reduced data collection procedures in the bDC/TMD, compared to the DC/TMD, resulting in likely concordant diagnostic meaningfulness in the data. Despite our optimism, the panel explicitly acknowledged that the reduced number of items would need testing either with primary or secondary data to ascertain the new instrument’s reliability, sensitivity and specificity, and utility.

Despite the many international attempts to implement the DC/TMD beyond academic and pain-specialist care settings, the overall use of the DC/TMD in non-specialist dental settings must today be considered as more or less non-existent. The bDC/TMD attempts to address the three major obstacles for implementation: i) the amount of training necessary to adequately use DC/TMD; ii) the belief that clinical implementation of the physical examination must be excessively time-consuming; and iii) the complexity of implementing the psychosocial axis. The bDC/TMD takes relies on ordinary language for the examination and can be self-taught, requires less time to complete, and has a simple set of instruments for the psychosocial axis; in addition, the bDC/TMD is likely to retain sufficient sensitivity and specificity for the most prevalent TMDs diagnoses. This should substantially improve the possibilities to implement bDC/TMD in non-specialist settings, for the benefit of patients and society since more patients should be identified, diagnosed, and treated earlier in the course of the disorder which improves prognosis ^34–36^.

The authors recommend self-management as the first step in managing any diagnosis made using the bDC/TMD ^21^ and suggest if this is implemented, patients are supported with this and then reviewed at 4-8 weeks to identify any improvement. To help reduce uncertainty of dentists over this type of management ^37^ several resources have been placed online (http://bit.ly/42zbTA6). If there is no improvement, or indeed a deterioration in the patient’s complaint, then a further specialist opinion on the diagnosis/management may be advisable at this stage. This is consistent with other guidelines for acute musculoskeletal pain with the propensity to become persistent ^38^.

The implementation of the bDC/TMD in non-specialist settings is outside the scope of this study but it likely requires a structured approach through implementation research. As an example, an implementation project in Sweden of an early version of the bDC/TMD comprised 107 general dentistry clinics and more than 2000 dental staff; the project used implementation principles of structured education and adaptation discussions for each clinic, video lectures, support from orofacial pain specialists, and follow-up. To date, the results have demonstrated an increase in TMDs relevant examinations by 46% and initiation of treatment by 31% (unpublished data, Alstergren).

The participants in the London workshop only included two non-specialists; however, their contributions and in particular critical disagreements aligned with experiences and general knowledge possessed by the other participants. The aim of the workshop was to find a basis for a set of procedures that can be used in non-specialist settings for the evaluation of TMDs, and all participants have experience with the general dentistry setting. Future research will necessarily incorporate critical feedback from stakeholders within general dental settings and pain non-specialist settings, for future revisions of the bDC/TMD.

## Conclusion

This study provides a Brief DC/TMD for use in non-specialist settings. The bDC/TMD was developed using a multidisciplinary and international process for reducing the published DC/TMD to bDC/TMD. This bDC/TMD can now be disseminated to non-specialist settings in which the bDC/TMD will be used to investigate validity and to improve the criteria for future versions.

## Supporting information

Supplemental information - bDC/TMD instrument

Supplemental information - Decision trees for instrument

## Data Availability

No data available

