## Supplemental information - bDC/TMD instrument for "Constructing the brief Diagnostic Criteria for Temporomandibular Disorders (bDC/TMD)"

**Self-report Section A - Pain Drawing**

Indicate the location of ALL of your different pains by shading in the area, using the diagrams that are most relevant. If there is an exact spot where the pain is located, indicate with a solid dot (●). If your pain moves from one location to another, use arrows to show the path.

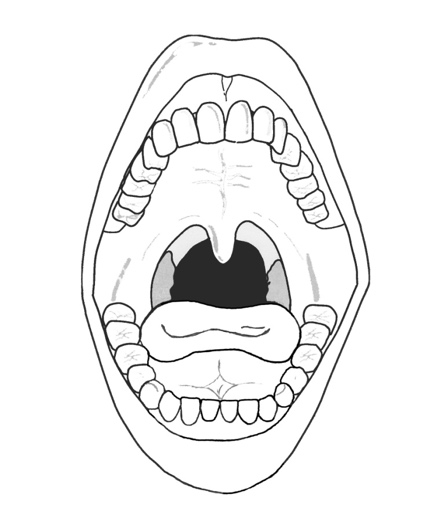

L

R

L

R

Right

Left

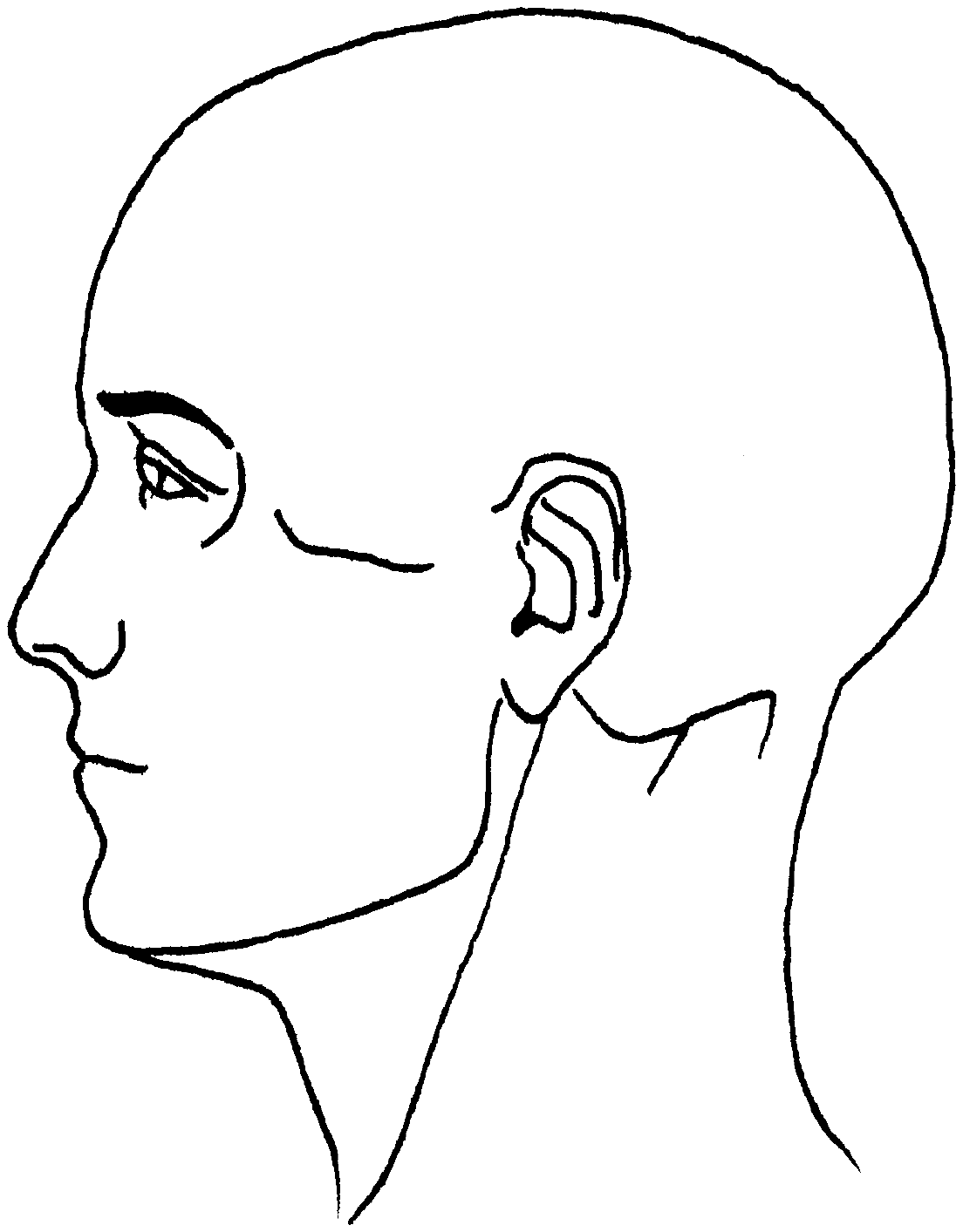

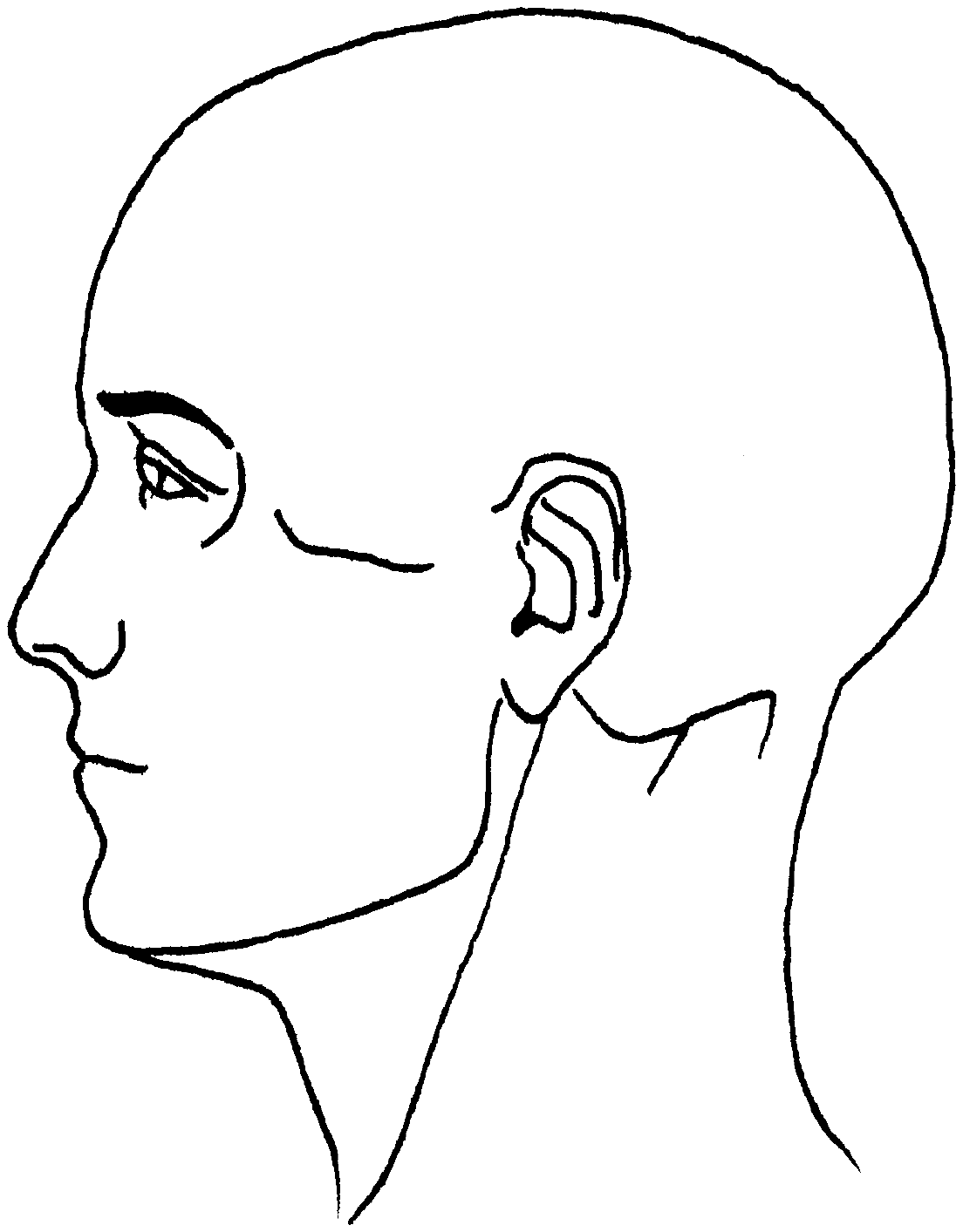

Right face

Left face

**Self-report Section B – Graded Chronic Pain Scale – 30 days**

| 1. How would you rate your facial pain RIGHT NOW? | | | | | | | | | | |
| --- | --- | --- | --- | --- | --- | --- | --- | --- | --- | --- |
| 0 | 1 | 2 | 3 | 4 | 5 | 6 | 7 | 8 | 9 | 10 |
| No pain | |  |  |  |  |  |  |  | Pain as bad  as could be | |
| 2. In the LAST 30 DAYS, how would you rate your WORST facial pain? | | | | | | | | | | |
| 0 | 1 | 2 | 3 | 4 | 5 | 6 | 7 | 8 | 9 | 10 |
| No pain | |  |  |  |  |  |  |  | Pain as bad  as could be | |
| 3. In the LAST 30 DAYS, ON AVERAGE, how would you rate your facial pain? | | | | | | | | | | |
| 0 | 1 | 2 | 3 | 4 | 5 | 6 | 7 | 8 | 9 | 10 |
| No pain | |  |  |  |  |  |  |  | Pain as bad  as could be | |
| [(Q1+Q2+Q3)/3]x10 = CPI = | | | | | | | | | | |
| 4. In the LAST 30 DAYS, how much has facial pain interfered with your DAILY ACTIVITIES? | | | | | | | | | | |
| 0 | 1 | 2 | 3 | 4 | 5 | 6 | 7 | 8 | 9 | 10 |
| No interference | |  |  |  |  |  |  |  | Unable to carry  on any activities | |
| 5. In the LAST 30 DAYS, how much has facial pain interfered with your RECREATIONAL, SOCIAL AND FAMILY ACTIVITIES? | | | | | | | | | | |
| 0 | 1 | 2 | 3 | 4 | 5 | 6 | 7 | 8 | 9 | 10 |
| No interference | |  |  |  |  |  |  |  | Unable to carry  on any activities | |
| 6. In the LAST 30 DAYS, how much has facial pain interfered with your ABILITY TO WORK, including housework? | | | | | | | | | | |
| 0 | 1 | 2 | 3 | 4 | 5 | 6 | 7 | 8 | 9 | 10 |
| No interference | |  |  |  |  |  |  |  | Unable to carry  on any activities | |
| [(Q4+Q5+Q6)/3]x10 = Disability = | | | | | | | | | | |

**Self-report Section C - PHQ 4 examining the effects of your pain**

Over the **PAST 2 WEEKS** have you been bothered by these problems? Please cross **ONLY one box** in each of 1-4

|  | ^NOT AT ALL^  ^(0)^ | ^SEVERAL DAYS (1)^ | ^MORE THAN HALF THE DAYS (2)^ | ^NEARLY EVERY DAY^  ^(3)^ |
| --- | --- | --- | --- | --- |
| 1) Feeling nervous, anxious, or on edge | **☐** | **☐** | **☐** | **☐** |
| 2) Not being able to stop or control worrying | **☐** | **☐** | **☐** | **☐** |
| 3) Feeling down, depressed, or hopeless | **☐** | **☐** | **☐** | **☐** |
| 4) Little interest or pleasure in doing things | **☐** | **☐** | **☐** | **☐** |

| If you checked off any problems, how difficult have these problems made it for you to do your work, take care of things at home, or get along with other people? | | | | | | | |
| --- | --- | --- | --- | --- | --- | --- | --- |
|  | Not difficult at all  ☐ |  | Somewhat difficult  ☐ |  | Very  difficult  ☐ |  | Extremely difficult  ☐ |

**Scoring and interpretation of self-report Axis 2 instrumentation**

**Pain diagram**

Wide-spread shading of the body mannikin in addition to any shading of the face/mouth should be explored with the patient and if this has not been investigated by other medical specialities the patient should be referred or asked to consult with their general medical practitioner for further evaluation.

**GCPS**

Calculations for characteristic pain intensity (CPI) and disability score are given under each sub-section. While individual item scores are useful guides during an interview, only computed scores should be relied on in terms of reliability and overall ease of interpretation.

Interpretation is that higher scores represent high pain intensity (CPI) or higher levels of interference (disability score).

An appreciation of the scores’ meaning can use a ‘thermometer type approach’. Note, however, considerable inter-individual variation occurs due to both the subjective nature of pain and its reporting as well as the many psychosocial factors that affect pain processing {Boonstra et al., 2016, #4685}:

| **Scoring** | 0 | 10 | 20 | 30 | 40 | 50 | 60 | 70 | 80 | 90 | 100 |
| --- | --- | --- | --- | --- | --- | --- | --- | --- | --- | --- | --- |
| **Interpretation of pain intensity (CPI) and interference** | None | Mild | | | | | Moderate | | Severe | | |

Based on available evidence, if both the CPI and interference scores are within the red zone it is more likely that prognosis may be affected, but this does not mean that simpler treatments should be omitted at the beginning of management.

**PHQ-4**

Scoring for each response is given in parenthesis below the frequency statement in the headings of columns 2-5. These are summed and interpretation can again be considered in a ‘thermometer type approach’ with ≥6 considered a yellow flag for review and ≥9 considered a red flag for further investigation and follow-up with more specialist services {Kroenke et al., 2009, #12990}:

| **Grade** | 0 | 1 | 2 | 3 | 4 | 5 | 6 | 7 | 8 | 9 | 10 | 11 | 12 |
| --- | --- | --- | --- | --- | --- | --- | --- | --- | --- | --- | --- | --- | --- |
| **Interpretation** | Normal | | | Mild | | | Moderate | | | Severe | | | |

The overall assessment of difficulty has no formal interpretation; rather, it serves as a discussion point with the patient. For example, the patient’s score is in the mild range but the patient reports “very difficult”; the clinician enquires into the nature of the difficulty and if that in any way affects their pain or coping with the pain. Similarly, the patient’s score is in the severe range but reports “somewhat difficult”; the clinician notes the reporting of severe symptoms but that the patient seems to be managing it well, and then enquires into how able to do that so effectively. The overall goal with the PHQ-4 (or any instrument substituted for it) is to make a determination regarding whether the patient is safe in terms of self-harm in relation to any reported distress, whether the reported distress is an important consideration for the patient’s reported pain intensity and exacerbations, and whether the patient needs further consultation and treatment by a mental health specialist.

**Self-report Section D – Symptom Questionnaire**

Reviewed by attending clinician at history taking during clinic appointment.

| **Item number** |  |  |  |
| --- | --- | --- | --- |
| **SQ1** | Have you ever had pain in your jaw, temple, in the ear, or in front of the ear on either side?  **If the answer is NO, please skip to question SQ5** | **No** ☐ | **Yes** ☐ |
| **SQ2** | How many years or months ago did your pain in the jaw, temple, in the ear, or in front of the ear first begin?  Years Months | | |

| **SQ3** | In the last 30 days, which of the following best describes any pain in your jaw, temple, in the ear, or in front of the ear on either side? Select ONE response | | |
| --- | --- | --- | --- |
|  | a) No pain | ☐ | |
|  | b) Pain comes and goes | ☐ | |
|  | c) Pain is always present | ☐ | |
|  | **If the answer is NO, please skip to question SQ5** | | |
| **SQ4** | In the last 30 days, did the following activities change any pain (that is, make it better or it worse) in your jaw or temple area on either side? | | |
|  | Chewing hard or tough food? | **No (0)**  ☐ | **Yes (1)**  ☐ |
|  | Opening your mouth or moving your jaw forward or to the side? | ☐ | ☐ |
|  | Jaw habits such as holding teeth together, clenching, grinding, or chewing gum? | ☐ | ☐ |
|  | Other jaw activities such as talking, kissing, or yawning? | ☐ | ☐ |

| **SQ5** | In the last 30 days, have you had any headaches that included the temple areas of your head? | **No** ☐ | **Yes** ☐ |
| --- | --- | --- | --- |
| **SQ6** | How many years or months ago did your temple headache first begin?  Years Months | | |
| **SQ7** | In the last 30 days, did the following activities change any headache (that is, make it better or make it worse) in your temple area on either side? | | |
|  | Chewing hard or tough food? | **No**  ☐ | **Yes**  ☐ |
|  | Opening your mouth or moving your jaw forward or to the side? | ☐ | ☐ |
|  | Jaw habits such as holding teeth together, clenching, grinding, or chewing gum? | ☐ | ☐ |
|  | Other jaw activities such as talking, kissing, or yawning? | ☐ | ☐ |

| **SQ8** | In the last 30 days, have you had any jaw joint noise(s) when you moved or used your jaw? | ☐ | ☐ |
| --- | --- | --- | --- |
| **SQ9** | Have you ever had your jaw lock or catch, even for a moment, so that it would not open ALL THE WAY?  **If answer No to this question skip to question SQ10** | ☐ | ☐ |
| **SQ10** | Was your jaw lock or catch severe enough to limit your jaw opening and interfere with your ability to eat? | ☐ | ☐ |
| **SQ11** | In the last 30 days, did your jaw lock so you could not open ALL THE WAY, even for a moment, and then unlock so you could open ALL THE WAY?  **If you answered NO to Question 11 then skip to Question 13.** | ☐ | ☐ |
| **SQ12** | Is your jaw currently locked or limited so that your jaw will not open ALL THE WAY? | ☐ | ☐ |
| **SQ13** | In the last 30 days, when you opened your mouth wide, did your jaw lock or catch even for a moment such that you could not close it from this wide-open position? | ☐ | ☐ |
| **SQ14** | In the last 30 days, when your jaw locked or caught wide open, did you have to do something to get it to close including resting, moving, pushing, or maneuvering it? | ☐ | ☐ |

**Additional information on headache (related to SQ5):**

Practitioners should note the red flags for headache below {Dodick, 2003, #12715} and refer urgently to neurology if these are present or refer as appropriate to the general medical practitioner in the cases of comorbid headache with TMD without any red flags.

Red flags SNOOP {Dodick, 2003, #12715}: **S**ystemic symptoms (fever, weight loss) or **S**econdary risk factors (HIV, malignancy); **N**eurological symptoms (confusion, impaired consciousness, cranial nerve pathology, motor or sensory disturbance); **O**nset sudden and abrupt (e.g., “thunderclap”); **O**lder (new and progressively worsening headache in age≥50); **P**rogressive and **P**revious history (headache worsening, changing in symptomatology, frequency or severity).

Migraine can effectively be screened for by use of the 3 questions below {Kim and Kim, 2006, #24153; Lipton et al., 2003, #56769}. Two or more affirmative responses to the questions below suggest a potential migrainous element to the headache and patients can be referred to their general medical practitioner for further investigation:

During the last 3 months, did you have the following with your headaches:

1. You felt nauseated or sick to your stomach? Yes/No
2. Light bothered you (a lot more than when you do not have headache)? Yes/No
3. Your headaches limited your ability to work, study, or do what you needed to do for at least one day? Yes/No
