## Supplemental information - Decision trees for instrument for "Constructing the brief Diagnostic Criteria for Temporomandibular Disorders (bDC/TMD)"

### Slide 1
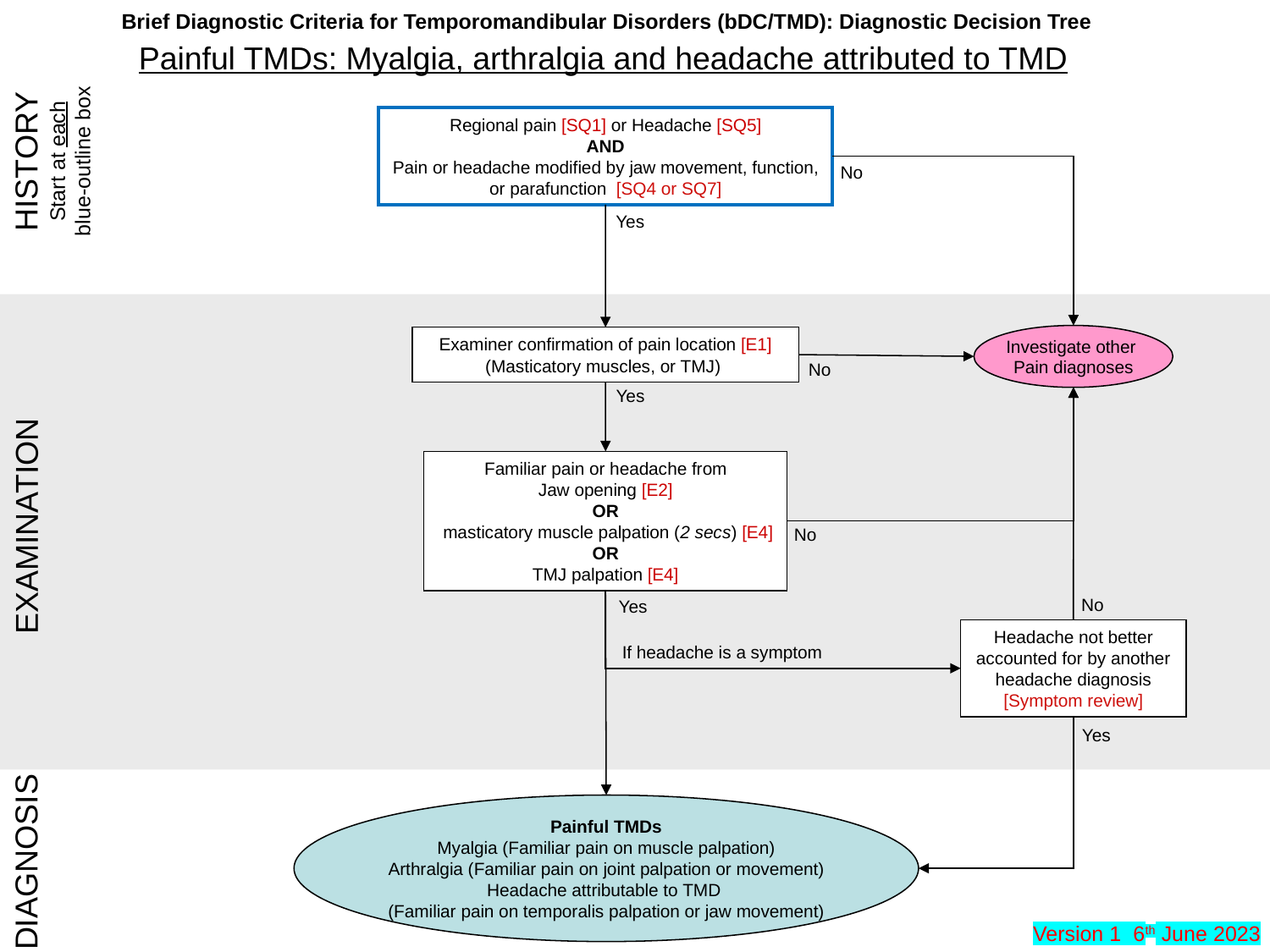

Brief Diagnostic Criteria for Temporomandibular Disorders (bDC/TMD): Diagnostic Decision Tree
Painful TMDs: Myalgia, arthralgia and headache attributed to TMD
Regional pain [SQ1] or Headache [SQ5]
AND
Pain or headache modified by jaw movement, function, or parafunction [SQ4 or SQ7]
HISTORY
Start at each
blue-outline box
No
Yes
Investigate other
Pain diagnoses
Examiner confirmation of pain location [E1]
(Masticatory muscles, or TMJ)
No
Yes
Familiar pain or headache from
Jaw opening [E2]
OR
 masticatory muscle palpation (2 secs) [E4]
OR
TMJ palpation [E4]
EXAMINATION
No
No
Yes
Headache not better accounted for by another headache diagnosis
[Symptom review]
If headache is a symptom
Yes
Painful TMDs
Myalgia (Familiar pain on muscle palpation)
Arthralgia (Familiar pain on joint palpation or movement)
Headache attributable to TMD
(Familiar pain on temporalis palpation or jaw movement)
DIAGNOSIS
Version 1 6^th^ June 2023

### Slide 2
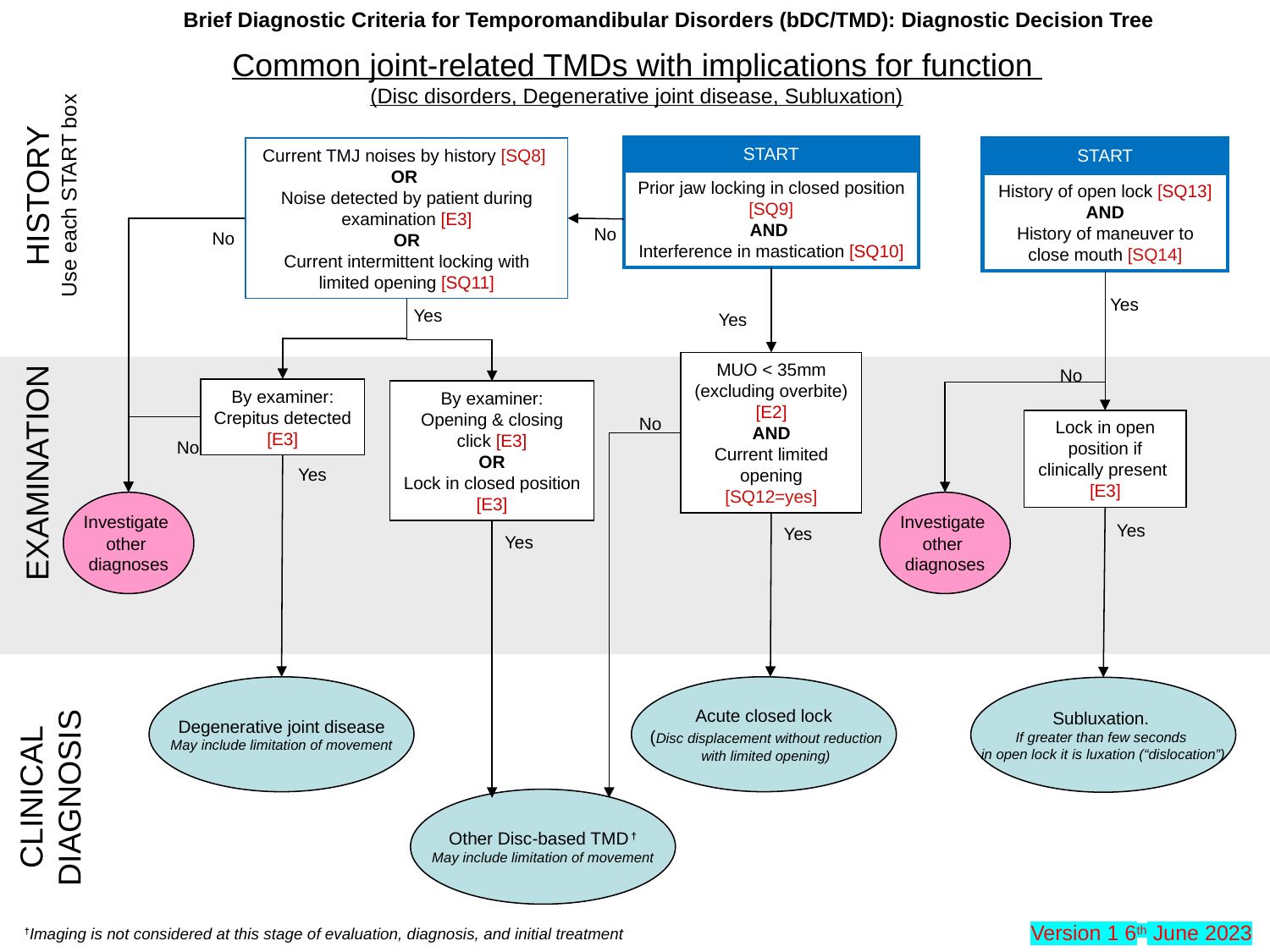

Brief Diagnostic Criteria for Temporomandibular Disorders (bDC/TMD): Diagnostic Decision Tree
Common joint-related TMDs with implications for function
(Disc disorders, Degenerative joint disease, Subluxation)
START
START
Current TMJ noises by history [SQ8]
OR
Noise detected by patient during examination [E3]
OR
Current intermittent locking with limited opening [SQ11]
HISTORY
Use each START box
Prior jaw locking in closed position [SQ9]
AND
Interference in mastication [SQ10]
History of open lock [SQ13] AND
History of maneuver to close mouth [SQ14]
No
No
Yes
Yes
Yes
MUO < 35mm (excluding overbite)
[E2]
AND
Current limited opening [SQ12=yes]
No
By examiner:
Crepitus detected
[E3]
By examiner:
Opening & closing click [E3]
OR
Lock in closed position [E3]
No
Lock in open position if clinically present
[E3]
No
EXAMINATION
Yes
Investigate
other
diagnoses
Investigate
other
diagnoses
Yes
Yes
Yes
Degenerative joint disease
May include limitation of movement
Acute closed lock
 (Disc displacement without reduction
 with limited opening)
Subluxation.
If greater than few seconds
in open lock it is luxation (“dislocation”)
CLINICAL DIAGNOSIS
Other Disc-based TMD ^†^
May include limitation of movement
Version 1 6^th^ June 2023
 ^†^Imaging is not considered at this stage of evaluation, diagnosis, and initial treatment
