## Supplemental information - Table 1 and Figure 1 for "Constructing the brief Diagnostic Criteria for Temporomandibular Disorders (bDC/TMD)"

**Supplementary information**

**Table 1 – List of Delphi participants and their affiliations**

|  | **Name** | | **Affiliation** | **Expertise** |
| --- | --- | --- | --- | --- |
|  | **Principal investigators** | | | |
|  | Justin | Durham | Newcastle University, UK | Oral surgery & orofacial pain |
|  | Richard | Ohrbach | University at Buffalo School of Dental Medicine, USA | Psychology and Orofacial pain |
|  | Per | Alstergren | Malmö University, Sweden | Orofacial pain |
|  | **Other Delphi participants** | | | |
|  | Lene | Baad-Hansen | Aarhus University, Denmark | Orofacial pain |
|  | Stephen | Davies | University of Manchester, UK | General & restorative dentistry |
|  | Anton | De Laat | KU Leuven, Belgium | Orofacial pain |
|  | Daniela | Gonclaves | São Paulo State University (Unesp), Brazil | Orofacial pain |
|  | Valeria | Gordon | University of Florida, USA | General & restorative dentistry |
|  | Jean-Paul | Goulet | Laval University, Canada | Orofacial pain |
|  | Birgitta | Henrikson | Malmö University, Sweden | Orofacial pain |
|  | Mick | Horton | College of General Dentistry, UK | General dentistry |
|  | Michail | Koutris | ACTA, University of Amsterdam and Vrije Universiteit Amsterdam, Netherlands | Orofacial pain |
|  | Alan | Law | University of Minnesota, USA | Endodontics, Orofacial pain |
|  | Thomas | List | Malmö University, Sweden | Orofacial pain |
|  | Frank | Lobbezoo | ACTA, University of Amsterdam and Vrije Universiteit Amsterdam, Netherlands | Orofacial pain |
|  | Ambra | Michelotti | University of Naples Federico II, Italy | Orthodontics, Orofacial pain |
|  | Don | Nixdorf | University of Minnesota, USA | Orofacial pain |
|  | Juan Fernando | Oyarzo | Universidad Andres Bello, Chile | Orofacial pain |
|  | Chris | Peck | University of Sydney, Australia | Orofacial pain |
|  | Chris | Penlington | Newcastle University, UK | Clinical Psychology and Orofacial pain |
|  | Karen | Raphael | New York University, USA | Clinical Psychology and Orofacial pain |
|  | Vivian | Santiago | New York University, USA | Orofacial pain |
|  | Sonia | Sharma | University at Buffalo School of Dental Medicine, USA | Orofacial pain |
|  | Peter | Svensson | Aarhus University, Denmark | Orofacial pain |
|  | Corine | Visscher | ACTA, University of Amsterdam and Vrije Universiteit Amsterdam, Netherlands | Physiotherapy and orofacial pain |
|  | Imamura | Yoshiki | Nihon University, Japan | Orofacial pain |

**Figure 1 – Gantt chart demonstrating work progress and pandemic delay**

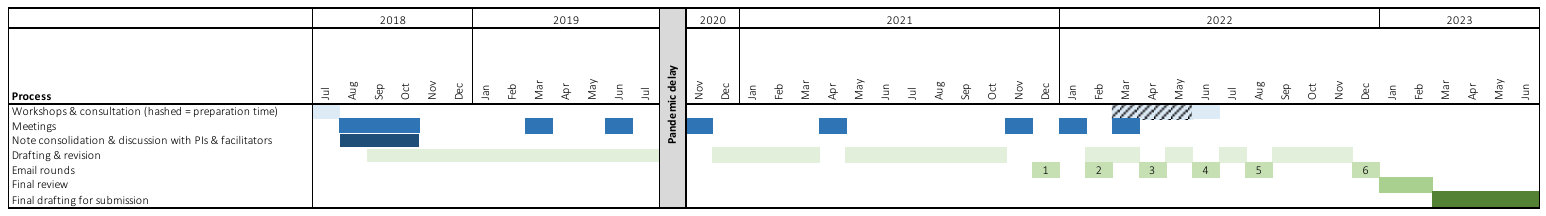
